# The Rise of Brazil’s Primary Care Digitalization: 12 Billion Records Across 27 Federative Units as a Foundation for Real-World Evidence and Scientific Democratization

**DOI:** 10.64898/2026.07.01.26356080

**Authors:** Pedro Marton Pereira, Alysson Nathan Girotto, Gabriela Machado Silva, Gustavo Duregger

## Abstract

**Background:** Brazil operates one of the world’s largest national primary health care information systems. Since 2013, the Health Information System for Primary Care (SISAB), the digital platform of the e-SUS primary care strategy, has collected standardized records of all clinical and community health activities performed by Family Health Strategy teams across all Brazilian municipalities. Yet comprehensive longitudinal analyses of this national data infrastructure remain scarce in the international literature.

**Methods:** Descriptive ecological study with time-series analysis using publicly available secondary data extracted from SISAB on May 18, 2026. Annual records were collected for four production types (Individual Care, Dental Care, Procedures, and Home Visits), disaggregated by all 27 Federative Units, covering April 2013 through March 2026. Per capita indicators were calculated using the 2022 Brazilian Demographic Census population by state.

**Results:** The cumulative total of SISAB records from April 2013 through March 2026 is 12,421,073,299. Annual volume grew from 53.3 million in 2014 (first full year) to 1.96 billion in 2025, an approximately 37-fold increase over 11 years. A 9.0% decline in 2020 was associated with the COVID-19 pandemic, with full recovery by 2021. Procedures rose from 21.6% to 35.9% of total records between 2014 and 2025, reflecting deepening clinical record completeness. Per capita records varied 3.2-fold across federative units, from 29.0 (Distrito Federal) to 92.9 (Tocantins). Home visits, performed predominantly by Community Health Workers, accounted for 41.9% of all records, with the highest per capita rates concentrated in the Northeast region.

**Conclusions:** SISAB constitutes a longitudinal national data infrastructure of exceptional scale, covering populations historically underrepresented in biomedical research: rural, Amazonian, and peri-urban communities. With over 12 years of continuous data and 184 million active registered patients, this system represents a strategic foundation for real-world evidence generation and the democratization of health science. Realizing this potential requires investment in data quality, interoperability, ethical governance, and scientific capacity aligned with Brazilian General Data Protection Law (Law nº 13,709/2018).

## INTRODUCTION

Primary Health Care (PHC) constitutes the first level of care in health systems, characterized by actions of health promotion and protection, disease prevention, diagnosis, treatment, rehabilitation, harm reduction, and health maintenance, with the aim of developing comprehensive care capable of positively impacting population health^1^.

In Brazil, PHC represents the main entry point of the Unified Health System (*Sistema Único de Saúde*, SUS) and the organizing center of the Health Care Network (*Rede de Atenção à Saúde*, RAS), guided by the principles of universality, accessibility, continuity of care, comprehensiveness, accountability, humanization, and equity^1^. Its priority care model is the Family Health Strategy (*Estratégia Saúde da Família*, ESF), an innovative model that places health at the center of the needs of individuals, families, and territories, playing a crucial role in the reorganization of PHC in Brazil, aligned with the principles of the SUS^2^.

Recognized as a priority by the Ministry of Health and by state and municipal managers, the ESF has driven the expansion, qualification, and consolidation of Primary Health Care, promoting a reorientation of work processes with great potential to strengthen care, expand problem-solving capacity, and generate positive impacts on the health of individuals and communities^2^. The minimum composition of each Family Health team (*equipe de Saúde da Família*, eSF) includes a physician, a nurse, a nursing assistant or technician, and community health workers (*Agentes Comunitários de Saúde*, ACS)^3^.

Despite this scale, Brazil’s PHC system has long been a “known unknown” in the global health literature: widely referenced as a model for low-and middle-income countries, yet rarely analyzed using its own national data. A central reason is that the information systems underpinning the ESF were, for most of its history, paper-based or fragmented across municipalities.

The international recognition of Brazil’s PHC model arrived before its digital transformation. The 2008 World Health Report^4^ highlighted the Family Health Strategy as an example of a middle-income country that successfully reoriented its health system toward primary care, with encouraging early results in health outcomes and patient satisfaction^4^.

A more recent assessment confirmed this international influence, documenting how four countries, Angola, Belgium, South Africa, and the United Kingdom, adapted elements of the ESF to strengthen their own health systems, with ACS emerging as the central transferable element in each case^5^.

This international recognition preceded the digital transformation of Brazilian PHC and was built upon a substantial workforce already active in the field. In the period prior to mandatory Health Information System for Primary Care (*Sistema de Informação em Saúde para a Atenção Básica*, SISAB) implementation, PHC had an average of 265,000 ACS, 41,000 physicians, and 45,800 nurses distributed across the national territory^6^, responsible for a monthly average of 10.9 million physician consultations, 7.56 million nursing consultations, and 28.9 million home visits, all recorded on paper forms and subsequently entered manually into the Primary Care Information System (*Sistema de Informação da Atenção Básica*, SIAB)^6^. The average PHC coverage during this period reached 69.5% of the Brazilian population^6^. This establishes an important baseline: the Brazilian PHC model had already achieved population-level impact on a national scale before any digital infrastructure for individual-level record keeping existed.

In the Americas, a 2024 Pan American Health Organization (PAHO) report assessed progress toward universal health coverage, analyzing indicators of unmet health care needs, essential services coverage, and financial protection. The report found that the essential services coverage index in the Region grew from 66 points in 2000 to 80 in 2019, reaching 74% in 2021, one of the highest among all WHO regions. In this context, Brazil is among a select group of countries, including Canada, Chile, Costa Rica, Cuba, the United States, and Uruguay, that achieved an essential services coverage index of 80% or above, representing only 20% of countries in the Region^7^.

The digitalization of this workforce-intensive system over the following decade positioned Brazil’s PHC data infrastructure as a relevant asset in the global movement toward Real-World Evidence (RWE). This movement, defined as the use of routinely collected health data to generate scientific evidence outside traditional clinical trials, has accelerated in recent years, driven by regulatory interest from agencies such as the U.S. Food and Drug Administration (FDA)^8^ and the European Medicines Agency (EMA)^9^, the COVID-19 pandemic, which exposed critical gaps in real-time health data infrastructure globally, and growing recognition that RWE can generate robust and complementary evidence to inform health decisions at population scale^10^. Brazil’s PHC data infrastructure sits at the intersection of these trends: large scale, diverse populations, long longitudinal span, and national coverage.

Prior work using Brazilian PHC data has demonstrated its potential: studies have used ACS-collected data to identify stroke survivors and map access to rehabilitation^11^, and to assess real-world statin use in cardiovascular prevention populations in primary care settings^12,13^. Yet no study has systematically characterized the full scale and evolution of the SISAB data infrastructure over its entire history.

This study aims to describe the healthcare production in Brazilian Primary Health Care by Federative Unit, using publicly available secondary data from SISAB, from April 2013 to March 2026.

## BACKGROUND

### The Brazilian primary health care system: a brief primer for international readers

Brazil is a federal republic of an estimated 213 million people, according to the Brazilian Institute of Geography and Statistics (IBGE)^14^ resident population estimate of July 1, 2025, across 8.5 million km^2^, comprising five macro-regions with profound socioeconomic and epidemiological heterogeneity. Its public health system, the *Sistema Único de Saúde* (SUS), guarantees universal access to health care as a constitutional right since 1988.

### From SIAB to SISAB: the digitalization arc

Brazil’s digitalization journey in PHC spans more than three decades. The roots lie in the early 1990s, when Brazil created the Community Health Agents Program Information System (*Sistema de Informação do Programa de Agentes Comunitários de Saúde*, SIPACS) to monitor the first Community Health Worker programs^15^. In 1998, this evolved into the SIAB^15^, the first national PHC information system, operational until 2015^6,16^. SIAB collected family-level aggregate data through paper forms completed by ACS, a landmark achievement for its time, but with structural limitations: it did not allow the identification of users or the coding of all diseases, and relied on manual consolidation of forms prone to delays and errors in the recording of care activities^6^.

The digital transformation effectively began in 2013, when the Ministry of Health established SISAB^17^ through Ordinance GM/MS nº 1.412, of July 10, 2013^18^, as the core of the e-SUS Primary Health Care strategy. In July 2025, the Ministry of Health established the Health Information System for Primary Health Care (*Sistema de Informação para a Atenção Primária à Saúde*, SIAPS) through Ordinance GM/MS nº 7.639, of July 18, 2025^19^, which amended the Consolidation Ordinance GM/MS nº 1, of September 28, 2017^20^. SIAPS is the result of an update to SISAB, designed to modernize the technological infrastructure and improve data management in PHC, making the handling of health information more agile and secure.

This movement was aligned with the National Primary Health Care Policy (*Política Nacional de Atenção Básica*, PNAB), established by Ordinance GM/MS nº 2.488, of October 21, 2011^21^, and updated by Ordinance GM/MS nº 2.436, of September 21, 2017^22^, which determines that all levels of government have a duty to develop, make available, and implement Primary Health Care information systems, ensuring mechanisms that guarantee the qualified use of these tools in health units, and that all professionals and municipal managers are responsible for entering, analyzing, and verifying the quality and consistency of data in these systems, using them for action planning^21,22^.

Unlike its predecessor, SISAB is based on individual-level care records that can be linked using national individual identifiers, such as the *Cartão Nacional de Saúde* (CNS) or the *Cadastro de Pessoas Físicas* (CPF). This data structure makes it possible, in subsequent analyses, to reconstruct individual care trajectories over time^6^. Mandatory national reporting became effective in January 2016 (Portaria nº 1.113/2015)^23^, initiating a steep adoption curve that accelerated progressively in subsequent years. Over more than a decade of operation, SISAB has reached a level of national scale and maturity that, to our knowledge, few PHC information systems in any country have achieved, making it a unique platform for population-level analysis of primary care production.

The Family Health Strategy organizes PHC through teams of approximately 3,500 residents per unit^24,25^. Each team includes a physician, a nurse, a nursing assistant or technician, and ACS. ACS are community members hired from the same territory they serve, a design that makes the ESF one of the few PHC models with systematic, doorstep-level contact with the entire registered population. As of April 2026, for an estimated population of 213,407,113, the team capacity reached 211,414,772 individuals, representing around 99.06% coverage^25^.

SISAB collects four types of production records: (1) Individual Care, comprising clinical consultations by any health professional; (2) Dental Care, comprising dental consultations and procedures; (3) Procedures, comprising clinical actions performed during or between consultations, such as blood pressure measurement, wound care, medication dispensing, and laboratory sample collection; and (4) Home Visits, comprising structured household visits conducted primarily by ACS, covering health assessment, guidance, referral, and social monitoring. Each record is timestamped and linked to a municipality and state, creating a georeferenced longitudinal database.

### What SISAB exposes nationally and what remains at the municipal level

SISAB in its national, publicly accessible form aggregates the four production types analyzed in this study. This is a deliberate design choice: the Ministry of Health collects these standardized production counts from all municipalities as the basis for federal financing and program monitoring. However, they represent a fraction of the data that actually flows through the e-SUS^26^ ecosystem.

The primary data source is the municipal level. Each of Brazil’s 5,570 municipalities operates one of two e-SUS^26^ data collection tools: the *Prontuário Eletrônico do Cidadão* (PEC, the electronic health record) or the simplified form-based system (CDS, *Coleta de Dados Simplificada*). The PEC generates structured clinical records that include sociodemographic data, household conditions, vaccination history, prescriptions, specialist referrals, laboratory results, and free-text clinical notes, constituting a substantially richer dataset than what is aggregated in national SISAB reports. As of December 2021, 19.3% of primary health care units in Brazil still used CDS, requiring manual data entry by health professionals^6^. Additionally, some municipalities may use third-party electronic health record systems rather than the PEC, but data transmission must occur through a municipal centralizer.

The national SISAB aggregate view therefore represents the nationally visible layer of a much larger distributed data infrastructure. State health secretariats have access to additional disaggregated data from their municipalities, and individual municipalities have full access to their own PEC records. Research partnerships at the local or regional level, between universities, research institutes, or health technology organizations and willing municipalities or state governments, can access individual-level records that would enable patient-level studies of exceptional richness and methodological depth. Such partnerships have already demonstrated feasibility, as illustrated by studies conducted with epHealth’s primary care database covering over two million patients^11,12^. All patient-level research requires submission to a Research Ethics Committee (*Comitê de Ética em Pesquisa*, CEP) and must comply with the Brazilian General Data Protection Law (*Lei Geral de Proteção de Dados Pessoais*, LGPD, Law nº 13,709/2018)^27^ provisions for sensitive health data, following established scientific and regulatory standards.

Data are publicly available at sisab.saude.gov.br and are, in the Ministry of Health’s own description, subject to revision, reflecting ongoing retroactive updates from municipalities. This is a known characteristic of administrative health data systems globally and is addressed in the limitations section.

## METHODS

### Study design

Descriptive ecological study with time-series analysis using publicly available secondary data. The unit of analysis is the SISAB production record, aggregated by year, record type, and Federative Unit. The protocol for this study is available on the Open Science Framework (OSF) at the following DOI: https://doi.org/10.17605/OSF.IO/M2S9F.

### Data source

Data were extracted from the SISAB public portal (sisab.saude.gov.br), module *Relatórios de Produção*, on May 18, 2026, using filters for all four production types, all federative units, and all available monthly competencies. The extraction covered April 2013, the first month of SISAB operation, through March 2026.

Active patient registrations (184,440,496) were obtained from the SISAB *Relatório de Cadastros*, with reference date April 2025, the most recent available before discontinuation of that specific report.

### Variables

Four production types were analyzed: Individual Care, Dental Care, Procedures, and Home Visits. Three analytical groupings were derived: (1) Direct clinical contact: Individual Care + Dental Care; (2) Extended PHC reach: Individual Care + Dental Care + Home Visits; (3) Total SISAB records: all four types combined.

### Per capita analysis

Per capita indicators were calculated by dividing cumulative records per state (April 2013 – March 2026) by the resident population from the 2022 Brazilian Demographic Census (IBGE)^24^. Five per capita indicators were calculated for each state: individual care, dental care, procedures, home visits, and total records per inhabitant.

### Statistical analysis

Data were organized into annual time series by production type and state. Year-over-year variation was calculated as percentage change relative to the previous year. A relative growth index (base year 2014 = 100) was computed to compare growth trajectories across record types. Analysis was performed in Python 3.12 using the pandas library (version 1.5+).

### Ethical considerations

This study uses exclusively public, aggregated, deidentified secondary data made available by the Brazilian Ministry of Health. No individual-level patient data were accessed. In accordance with Brazilian National Health Council Resolution nº 510/2016^29^ studies using only public secondary data without individual identification are exempt from ethics committee review. Any future studies using individual-level municipal records will require submission to a Research Ethics Committee (CEP) and full compliance with the Brazilian General Data Protection Law nº 13,709/2018)^27^.

## RESULTS

### Overall scale and historical trajectory

The cumulative total of SISAB records from April 2013 through March 2026 is **12**,**421**,**073**,**299**. Of these, 2,858,252,958 (23.0%) were Individual Care consultations, 443,998,974 (3.6%) Dental Care, 3,914,282,357 (31.5%) Procedures, and 5,204,539,010 (41.9%) Home Visits. Table 1 presents the complete annual series.

**Table 1.** Annual SISAB records by production type, Brazil, 2013–2026.

| Year | Individual Care | Dental Care | Procedures | Home Visits | Total |
| --- | --- | --- | --- | --- | --- |
| 2013* | 197,952 | 67,532 | 97,590 | 379,824 | **742,898** |
| 2014† | 14,042,696 | 3,670,886 | 11,514,943 | 24,077,117 | **53,305,642** |
| 2015 | 54,566,804 | 14,417,425 | 40,729,757 | 115,930,024 | **225,644,010** |
| 2016 | 132,122,450 | 36,906,516 | 108,401,125 | 283,081,193 | **560,511,284** |
| 2017 | 175,345,851 | 35,987,275 | 138,555,958 | 340,036,672 | **689,925,756** |
| 2018 | 223,092,598 | 43,169,856 | 208,975,198 | 363,338,927 | **838,576,579** |
| 2019 | 233,326,070 | 42,075,756 | 248,161,749 | 344,295,862 | **867,859,437** |
| 2020‡ | 199,424,062 | 19,822,831 | 233,861,823 | 336,987,279 | **790,095,995** |
| 2021 | 252,644,435 | 30,460,611 | 349,103,734 | 449,534,825 | **1,081,743,605** |
| 2022§ | 312,895,780 | 43,504,765 | 477,023,034 | 595,938,444 | **1,429,362,023** |
| 2023 | 356,713,145 | 50,408,187 | 580,092,789 | 668,280,921 | **1,655,495,042** |
| 2024 | 386,444,693 | 52,602,723 | 634,995,112 | 696,152,484 | **1,770,195,012** |
| 2025‖ | 414,551,864 | 56,875,660 | 703,292,001 | 784,895,827 | **1,959,615,352** |
| 2026<sup>¶</sup> | 102,884,558 | 14,028,951 | 179,477,544 | 201,609,611 | **498,000,664** |
| **Total** | **2,858,252,958** | **443,998,974** | **3,914,282,357** | **5,204,539,010** | **12,421,073,299** |

Source: SISAB/SAPS/Ministry of Health, extracted May 18, 2026.
\*2013: Apr–Dec (9 months); gradual rollout, not all federative units yet reporting. †2014: First full year. ‡2020: Start of the public health emergency declaration due to the COVID-19 pandemic in Brazil. §2022: End of the public health emergency declaration due to the COVID-19 pandemic in Brazil. ¶2025: Historic record. ¶2026: January–March only (partial year).

In 2014, the first full year of SISAB operation, 53.3 million records were registered. By 2025, this figure reached **1**,**959**,**615**,**352**, a 36.8-fold increase over 11 years, and the system’s historic annual record. The mean monthly volume in 2025 was 163.3 million records, equivalent to 5.4 million per day or more than 24 home visits per second throughout the year.

In the first three months of 2026, 498,000,664 records were registered, an annualized pace of approximately 1.99 billion, indicating continued expansion beyond the 2025 record.

### Shifts in record type composition

The relative composition of records shifted substantially over the study period. Procedures, which represented 21.6% of total records in 2014, grew to 35.9% in 2025, the largest relative increase of any category. Individual Care declined from 26.3% to 21.2% as a share of total, reflecting the proportionally faster growth of other categories rather than an absolute decline (Individual Care grew 29.5-fold in absolute terms between 2014 and 2025). Home Visits remained the largest single category throughout, at 41.9% of the cumulative total.

Dental Care showed the most pronounced COVID-19 sensitivity: its 2020 volume fell 52.9% relative to 2019, compared with a 9.0% overall decline. By 2021, total records had surpassed 2019 levels by 24.6%, with Dental Care recovering more slowly.

### Geographic variation: per capita analysis across 27 federative units

Per capita total records ranged 3.2-fold across federative units, from 29.0 (Distrito Federal) to 92.9 (Tocantins). Table 2 presents all 27 federative units ranked by cumulative per capita total.

**Table 2.** Cumulative SISAB records and per capita indicators by Federative Unit, Brazil, 2013–March 2026.

| # | State | Region | Pop.<br>2022* | Total<br>Records | pc<br>IC<sup>†</sup> | pc<br>DC<sup>‡</sup> | pc<br>Proc<sup>§</sup> | pc<br>HV | pc<br>Total<sup>¶</sup> |
| --- | --- | --- | --- | --- | --- | --- | --- | --- | --- |
| 1 | **Tocantins** | North | 1,607,363 | 149,278,447 | 18.3 | 3.1 | 30.1 | 41.4 | **92.9** |
| 2 | **Paraíba** | Northeast | 3,999,415 | 352,944,859 | 17.7 | 3.4 | 17.1 | 50.1 | **88.2** |
| 3 | **Santa Catarina** | South | 7,762,154 | 662,261,319 | 23.1 | 2.6 | 30.9 | 28.7 | **85.3** |
| 4 | **Alagoas** | Northeast | 3,127,683 | 265,649,589 | 15.7 | 3.2 | 18.2 | 47.8 | **84.9** |
| 5 | **Piauí** | Northeast | 3,270,174 | 268,600,969 | 15.1 | 2.9 | 17.1 | 47.1 | **82.1** |
| 6 | Mato Grosso do Sul | Central-West | 2,756,700 | 225,579,035 | 17.0 | 2.7 | 26.6 | 35.5 | 81.8 |
| 7 | Minas Gerais | Southeast | 20,732,660 | 1,541,172,849 | 17.6 | 2.5 | 20.2 | 34.0 | 74.3 |
| 8 | Paraná | South | 11,516,840 | 845,898,462 | 18.6 | 2.8 | 33.4 | 18.7 | 73.4 |
| 9 | Ceará | Northeast | 8,791,688 | 629,242,728 | 17.2 | 2.7 | 16.7 | 35.0 | 71.6 |
| 10 | Maranhão | Northeast | 6,775,152 | 482,324,834 | 10.4 | 1.9 | 10.7 | 48.1 | 71.2 |
| 11 | Espírito Santo | Southeast | 3,833,486 | 266,400,900 | 13.5 | 2.1 | 28.8 | 25.1 | 69.5 |
| 12 | Amazonas | North | 3,952,262 | 270,565,032 | 12.2 | 2.1 | 19.5 | 34.7 | 68.5 |
| 13 | Rio Gde do Norte | Northeast | 3,302,406 | 225,942,924 | 14.5 | 3.3 | 17.1 | 33.5 | 68.4 |
| 14 | Rio Grande do Sul | South | 10,880,749 | 743,195,223 | 19.2 | 2.2 | 30.0 | 16.9 | 68.3 |
| 15 | Sergipe | Northeast | 2,068,017 | 139,597,135 | 13.0 | 2.4 | 13.8 | 38.3 | 67.5 |
| 16 | Mato Grosso | Central-West | 3,658,813 | 235,790,755 | 14.0 | 2.0 | 23.7 | 24.8 | 64.4 |
| 17 | Pernambuco | Northeast | 9,058,155 | 562,808,007 | 11.2 | 2.5 | 11.7 | 36.6 | 62.1 |
| 18 | Bahia | Northeast | 14,141,626 | 817,334,516 | 10.0 | 2.1 | 13.1 | 32.6 | 57.8 |
| 19 | Roraima | North | 636,707 | 33,450,017 | 10.6 | 1.2 | 25.1 | 15.7 | 52.5 |
| 20 | Rondônia | North | 1,815,278 | 94,766,901 | 9.6 | 0.8 | 16.0 | 25.7 | 52.2 |
| 21 | Acre | North | 906,876 | 45,596,011 | 10.5 | 1.5 | 12.1 | 26.1 | 50.3 |
| 22 | Pará | North | 8,116,132 | 405,978,118 | 8.5 | 1.5 | 10.9 | 29.1 | 50.0 |
| 23 | Goiás | Central-West | 7,206,589 | 350,725,020 | 12.1 | 1.8 | 14.9 | 19.8 | 48.7 |
| 24 | Rio de<br>Janeiro | Southeast | 16,054,524 | 745,330,589 | 10.8 | 2.2 | 17.1 | 16.3 | 46.4 |
| 25 | São<br>Paulo | Southeast | 44,420,459 | 1,949,201,475 | 12.6 | 1.7 | 17.8 | 11.8 | 43.9 |
| 26 | Amapá | North | 829,494 | 29,805,703 | 6.0 | 1.4 | 8.4 | 20.1 | 35.9 |
| 27 | Distrito<br>Federal | Central-West | 2,817,381 | 81,631,882 | 8.5 | 1.2 | 16.8 | 2.4 | 29.0 |

Source: SISAB/SAPS/Ministry of Health, extracted May 18, 2026.
\*Population: 2022 IBGE Demographic Census<sup>28</sup>. <sup>†</sup>pc IC: per capita Individual Care. <sup>‡</sup>pc DC: per capita Dental Care. <sup>§</sup>pc Proc: per capita Procedures. <sup>||</sup>pc HV: per capita Home Visits. <sup>¶</sup>pc Total: sum of all types. Note: Sorted by pc Total descending.

The geographic pattern is not uniform across record types. For Home Visits per capita, the Northeast region dominates: Paraíba (50.1), Maranhão (48.1), Alagoas (47.8), and Piauí (47.1), all federative units with historically dense ACS networks and high ESF coverage. For Individual Care and Procedures per capita, the South region leads: Santa Catarina (23.1 individual care/capita, 30.9 procedures/capita), Rio Grande do Sul (19.2 and 30.0), and Paraná (18.6 and 33.4), reflecting earlier e-SUS adoption and greater clinical record completeness per consultation.

The Distrito Federal (federal capital) recorded the lowest per capita in every category, particularly Home Visits (2.4/capita), consistent with its more hospital-centric care model and proportionally lower ACS coverage relative to other federative units.

## DISCUSSION

### Scale in global context

To our knowledge, no comparable national PHC information system exists with this combination of scale, longitudinal depth, geographic coverage, and public accessibility. The UK’s CPRD (Clinical Practice Research Datalink) covers approximately 11.3 million patients but is not publicly accessible^30^. The Danish National Health Registers are often cited as a gold standard for record linkage but cover a population of 5.9 million^31^. Brazil’s SISAB, by contrast, covers 184 million active registrations across a continent-sized country with extreme socioeconomic diversity, and is available for download by any researcher.

The approximately 37-fold growth in annual records between 2014 and 2025 reflects two simultaneous processes: expansion of ESF coverage and progressive adoption of the e-SUS platform by municipalities. This growth trajectory mirrors broader patterns documented in low- and middle-income countries, where digital health initiatives have undergone substantial transformations propelled by technological advancements, increased internet accessibility, and progressive adoption of digital tools in health care delivery^32^, but at an unusual scale for a middle-income country, both in terms of population covered and the depth of its community health worker infrastructure.

### What the geographic heterogeneity reveals

The inverse relationship between Home Visit per capita, highest in the Northeast, and Procedure per capita, highest in the South (Table 2), reflects genuinely different PHC models rather than data quality differences alone. The Northeast’s ESF was built around ACS-intensive community engagement, a design rooted in the region’s rural geography and historically weaker health infrastructure, where physician density reaches only 1.93 per 1,000 inhabitants compared to 3.27 in the South^33^, reflecting deep and persistent regional inequalities in income distribution and access to essential health services^34^. The South’s higher procedure counts per capita reflect more complete clinical documentation, earlier e-SUS adoption, and a model with relatively greater physician density.

This heterogeneity is not a limitation of the data. It is the data’s primary scientific value. It means that analyses using SISAB can detect differences in care delivery models, ACS intensity, and clinical documentation practice across the full spectrum of Brazilian geography, including populations in the Amazon basin and semi-arid Northeast that are almost entirely absent from international clinical trial registries^35^.

### Toward scientific democratization

The concept of scientific democratization in health research encompasses two related ideas: making research methods and findings accessible across economic strata, and ensuring that the populations studied in science reflect the actual diversity of humanity. On both dimensions, SISAB offers an unusual opportunity.

The federative units with the highest SISAB records per capita are among the most socioeconomically vulnerable in Brazil, with high proportions of rural, indigenous, and Afro-Brazilian communities. According to the 2022 Brazilian Demographic Census, the North and Northeast together account for 75.7% of the country’s indigenous population, and the Northeast records the highest proportion of Black individuals of any Brazilian region^28^. Brazil’s large, diverse, and highly admixed population offers a fundamental yet often underrepresented cohort for global health research^10^, and these are precisely the populations systematically excluded from randomized controlled trials and pharmacoepidemiological studies that rely on hospital or insurance databases^31^. In this sense, SISAB is not merely a large dataset. It is a dataset that reaches where conventional research does not.

Prior studies using this infrastructure demonstrate the feasibility of RWE research at population scale, including real-world statin utilization in cardiovascular prevention populations and the identification of stroke survivors for rehabilitation access mapping^11,12^. Each of these studies would have been impossible without the ACS data collection infrastructure documented here.

### Conditions for realizing the potential

The existence of 12 billion records documented in this study (Table 1) does not automatically translate into research-grade evidence. Three structural challenges must be addressed.

#### Data quality and completeness

SISAB records are administrative in origin. The sharp rise in Procedures as a share of total records, from 13% in 2013 to 36% in 2025 (Table 1), may reflect both genuine expansion of clinical activity and improved documentation habits. Distinguishing these mechanisms requires linkage with other data sources, which is currently limited^6^. Record completeness remains heterogeneous across municipalities: federative units with longer e-SUS adoption histories (South, Southeast) systematically show higher per capita procedure counts than federative units with more recent adoption (parts of the North) (Table 2).

#### Interoperability and linkage

SISAB captures PHC production but does not link to hospital admissions, laboratory results, medication dispensing records, or mortality data at the individual level. Brazil has multiple parallel health information systems (SIH-SUS for hospitalizations, SINAN for notifiable diseases, SIM for mortality, BNAFAR for pharmacy) that, if linked, would constitute a research infrastructure comparable to the Nordic registers^31^. The national *Rede Nacional de Dados em Saúde* (RNDS) initiative is building this interoperability layer^10^, but linkage at research scale remains a work in progress.

#### Governance, ethics, and data protection

Brazil’s General Data Protection (LGPD, 13,709/2018)^27^, analogous to the EU’s GDPR^36^, governs the use of personal health data. While SISAB data used in this study are publicly available and aggregated, any individual-level research linkage requires ethical approval, data governance agreements, and compliance with LGPD provisions for sensitive health data. Robust governance frameworks, with patient representation, transparent data access protocols, and mechanisms ensuring benefit returns to the communities studied, are prerequisites for responsible research use at scale.

## CONCLUSION

Between April 2013 and March 2026, Brazil’s primary care information system accumulated 12,421,073,299 records, the product of 12 years of digital health infrastructure built inside one of the world’s most ambitious universal health systems. The nearly 37-fold growth in annual volume, the near-complete national geographic coverage, and the 184 million active patient registrations place SISAB among the largest and most accessible national PHC data infrastructures globally.

For the international health research community, SISAB represents an underutilized scientific resource. Its greatest value may lie not in its size but in its reach: the communities it captures, including Amazonian riverine populations, semi-arid northeastern smallholders, and peri-urban peripheries of large cities, are precisely those that conventional research has systematically failed to represent. Unlocking this potential requires not only analytical capacity, but governance frameworks that ensure the communities that generated these records are the primary beneficiaries of the knowledge they produce.

### Limitations

This study has several limitations that must be considered when interpreting these findings. First, SISAB national records reflect documented rather than actual production. Under-documentation is likely, particularly in the early years of the system, in the early years of the system, around 2013 to 2015, and in municipalities with weaker technological infrastructure, connectivity, or staff training in e-SUS. The steep growth in the early years partly reflects expanding adoption rather than expanding activity.

Second, the national SISAB portal provides access to only four aggregate production types. The full e-SUS ecosystem, including individual-level PEC records with diagnoses, prescriptions, laboratory results, vaccination histories, referrals, and sociodemographic profiles, is not accessible through the national public portal. SISAB is best understood as a national aggregator of a much richer distributed data infrastructure. The present analysis covers only the nationwide-visible layer; the clinical depth exists primarily at the municipal level.

Third, the transition from SIAB (aggregate, family-level) to SISAB (individual-level) and the phased municipal adoption create a structural break in the time series around 2013–2016. The growth observed in this period reflects both genuine expansion and system substitution effects, which cannot be fully disentangled from aggregate data alone. A prior Bayesian structural time-series analysis estimated that the SIAB-to-SISAB transition resulted in a measurable discontinuity in reported records^6^.

Fourth, data quality challenges at the municipal level, including duplicate patient registrations, inconsistent coding practices, connectivity gaps in rural areas, and varying staff capacity for clinical documentation, affect record completeness and comparability across municipalities. National SISAB figures reflect the sum of these heterogeneous quality environments. Certification and deduplication of patient registrations, standardization of coding practices, and investment in municipal data infrastructure are necessary conditions for using these data at research-grade level.

Fifth, the per capita denominators use total state population (IBGE 2022 Census)^28^ rather than the ESF-registered population, which may underestimate actual intensity in high-coverage federative units. Sixth, 2013 (9 months) and 2026 (3 months) are partial years and were excluded from trend analyses to avoid distortion. Finally, although municipalities using third-party electronic health record systems are expected to transmit production data through a municipal centralizer to SISAB, not all data may be captured due to potential integration limitations, meaning that some clinical detail may be retained locally and fall outside the scope of federal aggregation.

## SUPPLEMENTARY INFORMATION

### Data availability

All data used in this study are publicly available at sisab.saude.gov.br. Those interested in additional details regarding the analytical procedures may contact the corresponding author.

### Funding

This study received no external funding.

### Authors’ contributions

1. Pedro Marton Pereira: conceptualization, methodology, data curation, formal analysis, writing – original draft, and final approval.
2. Alysson Nathan Girotto: conceptualization, supervision, writing – review & editing, and final approval.
3. Gabriela Machado Silva: methodology, writing – original draft, writing – review & editing, and final approval.
4. Gustavo Duregger: formal analysis, writing – original draft, writing – review & editing, and final approval. All authors approved the final version of the manuscript and are accountable for all aspects of the work, ensuring the accuracy and integrity of any part of the study.

## Acknowledgements

The authors thank the health professionals of the Family Health Strategy teams across Brazil whose daily clinical work generated the records analyzed in this study, and the SAPS/Ministry of Health technical team for maintaining SISAB as a publicly accessible infrastructure.

## Artificial Intelligence use statement

During manuscript preparation, the generative AI tool Claude Sonnet 4.6 (Anthropic) was used for language editing, translation, and reference formatting. All AI-assisted content was reviewed and verified by the authors, who take full responsibility for the accuracy and integrity of the final manuscript.

